# Acute Myocarditis Complicated by Ventricular Arrhythmias: Prevalence, Outcomes and Acute Ablation Results

**DOI:** 10.64898/2026.03.02.26347476

**Authors:** Eran Leshem, Tzurit Kusniec, Adam Folman, Mark Kazatsker, Ofer Kobo, Ariel Roguin, Gilad Margolis

## Abstract

**Background:** Acute myocarditis is typically self-limiting and resolves spontaneously in most cases. However, ventricular arrhythmias (VA) complications, which may be life-threatening are associated with higher rates of in-hospital complications and mortality. Catheter ablation is occasionally required for acute myocarditis associated ventricular tachycardia (VT), but data on its procedural use and outcomes, in this setting, remain limited. We aimed to determine the prevalence of VA among patients hospitalized for acute myocarditis and to evaluate the subset who underwent in-hospital VT ablation, including their acute outcomes.

**Methods:** Retrospective analyzed data from the National Inpatient Sample (NIS) database for U.S. hospitalizations with a diagnosis of myocarditis between 2016 and 2019. In-hospital outcomes were compared between patients with and without VA. Subgroup analysis examined patients with acute myocarditis associated VT stratified by whether VT ablation was performed. Patient demographics, comorbidities, procedures, and outcomes were identified using ICD-10-CM codes.

**Results:** Among an estimated 17,845 hospitalizations for acute myocarditis, 8.4% (n=1,505) had VA (including 7.7% with VT). Patients with VA were more likely to have structural heart disease, renal disease, infectious etiologies, anemia, and atrial arrhythmias, despite lower prevalence of some traditional cardiac risk factors. VA was associated with markedly worse outcomes, including 5.5-fold higher in-hospital mortality (10% vs 1.6%; p<0.001). Multivariate analysis revealed that VA during admission for acute myocarditis was an independent significant risk factor for cardiac complications (aOR=4.8), total complications (aOR=4.2) and in hospital mortality (aOR=5.1) (p<0.001 for each analysis).

Among patients with VT, catheter ablation was performed in 13.7% (n=190), more commonly with infectious etiologies. Ablated patients, compared to those without ablation, experienced significantly higher rates of in-hospital complications (73.7% vs 42.3%; p<0.001) and mortality (15.8% vs 6.7%; p<0.001).

**Conclusions:** VA complicating acute myocarditis, portends significantly worse in-hospital outcomes. Although ablation was performed in approximately 1 in 7 patients with VT, those undergoing the procedure had less favorable acute results. Further prospective research is warranted to define optimal criteria for ablation and expected outcomes in this high-risk population.

## INTRODUCTION

Acute myocarditis is an inflammatory disease of the myocardium, triggered by a variety of etiologies including viral infections, autoimmune disorders, and exposure to cardiotoxic agents. Although many cases follow a benign and self-limiting course, a clinically significant subset may develop life-threatening arrhythmias.^1,2^ Previous studies have demonstrated an increase in incidence of arrhythmias with increasing age and comorbidities.^3^ Overall, ventricular arrhythmias (VA) occur in approximately 10% of patients with acute myocarditis, with ventricular fibrillation (VF) or cardiac arrest in about 3%. These events are associated with significantly higher risks of in-hospital complications and mortality.^4–6^

In myocarditis-related ventricular tachycardia (VT) refractory to antiarrhythmic drugs (AAD) therapy, catheter ablation was shown to be feasible. ^7,8^ Evidence from relatively small-scale studies suggests that ablation outcomes in the post-inflammatory stage of myocarditis are better than those during acute myocarditis.^9,10^ This may be explained by the more irregular and polymorphic VA observed in acute myocarditis, compared with the more regular and monomorphic VA typically seen in healed myocarditis.^11^ Current guidelines provide a Class IIa recommendation (Level of Evidence C) for catheter ablation in patients with post-myocarditis VA, whereas specific guidelines for the treatment of ventricular arrhythmias in acute myocarditis patients are lacking.^2,12^

The present study aims to identify and characterize patients on a national scale that were diagnosed with acute myocarditis and developed VA, and to evaluate the proportion of these patients who underwent catheter ablation during their hospitalization and their acute outcomes.

## METHODS

### Data source

Data for this study were obtained from the National Inpatient Sample (NIS), the Healthcare Cost and Utilization Project (HCUP), and Agency for Healthcare Research and Quality (AHRQ).^13^

The NIS is the largest publicly available all-payer database of inpatient hospitalizations in the United States, representing approximately a 20% stratified sample of discharges from U.S. hospitals. As the NIS comprises only de-identified data, this study was exempt from institutional review board by the local Human Research Committee.

The NIS provides comprehensive patient- and hospital-level data, including demographic characteristics, primary and secondary diagnoses and procedures, comorbid conditions, length of stay (LOS), hospital region, teaching status, bed size, and total hospitalization costs. To generate national estimates, researchers can apply sampling weights provided by HCUP at both the patient and hospital levels.

For the purposes of this analysis, data spanning the years 2016 to 2019 were extracted. Diagnoses and procedures were identified using International Classification of Diseases, 10th Revision, Clinical Modification (ICD-10-CM) codes. Each index hospitalization record includes one principal discharge diagnosis, up to 39 secondary diagnoses, and a maximum of 25 procedures.

### Study Population and Variables

The study included patients 18 years of age or older who had acute myocarditis as the primary discharge diagnosis using ICD-10-CM codes: I40, I400, I401, I408, I409, I41, I514, I012, I090, J1082, J1182, A381, A3952, B2682, B332, B3320, B3322, B3324, B5881, D8685.

ICD-10-CM codes I47.0, I47.2, I47.20, I49.01, I49.02 and I47.29 were used to identify patients with a diagnosis of VA. Patients undergoing percutaneous VT catheter ablation in the same hospital admission were identified with the following codes: 025K3ZZ, 025L3ZZ, 02BK3ZZ, 02BL3ZZ, 025M3ZZ, 02583ZZ. Flow chart of patient identification and inclusion is presented in figure 1.

**Figure 1:**
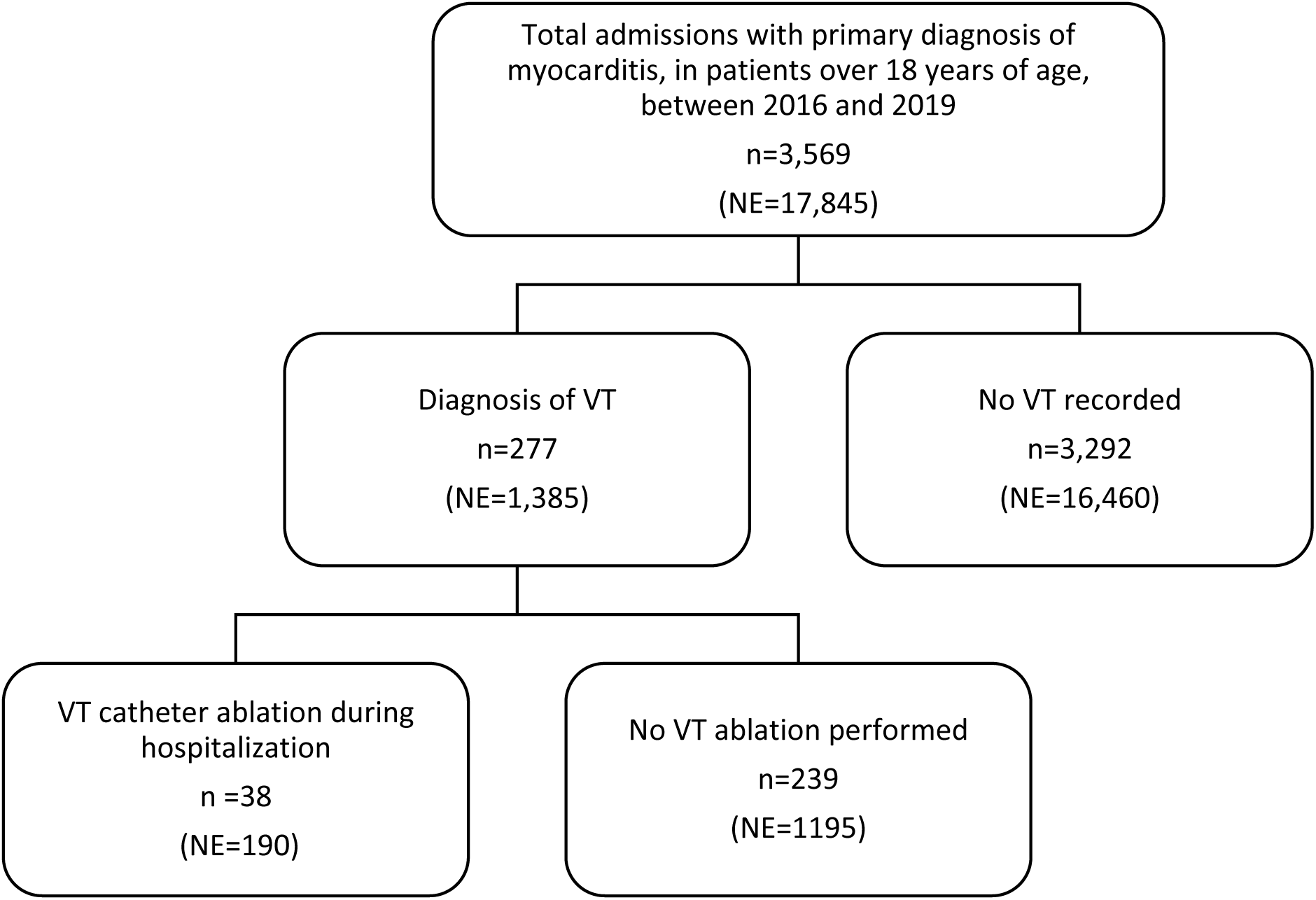
Flow chart of acute myocarditis patient identification and inclusion from the NIS registry. n=actual number of hospitalizations in the data set; NE=national estimate of hospitalizations; VT=ventricular tachycardia.

Patient demographics including age, sex, and race, were collected from the database. Patient-level data including comorbid conditions, secondary diagnoses, and relevant in-hospital outcomes and procedures were extracted from the database using ICD-10-CM/PCS codes (Appendix Table 1 and Appendix Table 2)

**Table 1:** Baseline characteristics acute myocarditis patients complicated by VA.

| Variable | Total Patients with<br>myocarditis<br>(NE=17845) | VA<br>(NE=1505) | No VA<br>(NE=16340) | p value |
| --- | --- | --- | --- | --- |
| Age (years) | 35 (22-53) | 36 (19-57) | 35 (22-53) | 0.193 |
| Female | 6150 (34.5%) | 490 (32.6%) | 5660 (34.6%) | 0.104 |
| White ethnicity | 10280 (60.5%) | 845 (59.5%) | 9435 (60.6%) | 0.438 |
| Diabetes mellitus | 1355 (7.6%) | 75 (5.0%) | 1280 (7.8%) | <0.001 |
| Hypertension | 5460 (30.6%) | 465 (30.9%) | 4995 (30.6%) | 0.792 |
| Dyslipidemia | 4030 (22.6%) | 260 (17.3%) | 3770 (23.1%) | <0.001 |
| Renal disease | 935 (5.2%) | 105 (7.0%) | 830 (5.1%) | 0.002 |
| Hepatic disease | 470 (2.6%) | 55 (3.7%) | 415 (2.6%) | 0.010 |
| Smoking | 5140 (28.8%) | 365 (24.3%) | 4775 (29.2%) | <0.001 |
| Obesity (BMI>30) | 1740 (9.8%) | 120 (8.0%) | 1620 (9.9%) | 0.015 |
| Valvular heart disease | 1485 (8.3%) | 175 (11.6%) | 1310 (8.0%) | <0.001 |
| Ischemic heart disease | 3875 (21.7%) | 315 (20.9%) | 3560 (21.8%) | 0.440 |
| Ischemic cardiomyopathy | 185 (1.0%) | 30 (2.0%) | 155 (0.9%) | <0.001 |
| Anemia | 510 (2.9%) | 60 (4.0%) | 450 (2.8%) | 0.006 |
| Alcohol abuse | 265 (1.5%) | 20 (1.3%) | 245 (1.5%) | 0.601 |
| Infectious disease* | 995 (5.6%) | 195 (13.0%) | 800 (4.9%) | <0.001 |
| Inflammatory disease** | 560 (3.1%) | 45 (3.0%) | 515 (3.2%) | 0.731 |
| CDMF-CCI score*** | 0 (0-1) | 1 (0-2) | 0 (0-1) | <0.001 |
| Income percentile |  |  |  |  |
| 0–25 | 4380 (24.9%) | 370 (24.8%) | 4010 (24.9%) | <0.001 |
| 26–50 | 4405 (25.1%) | 280 (18.8%) | 4125 (25.7%) |  |
| 51–75 | 4490 (25.6%) | 415 (27.9%) | 4075 (25.3%) |  |
| 76–100 | 4295 (24.4%) | 425 (28.5%) | 3870 (24.1%) |  |
| Hospital type |  |  |  |  |
| Rural | 710 (4.0%) | 40 (2.7%) | 670 (4.1%) | <0.001 |
| Urban, non-teaching | 3020 (16.9%) | 170 (11.3%) | 2850 (17.4%) |  |
| Urban, teaching | 14115 (79.1%) | 1295 (86.0%) | 12820 (78.5%) |  |

\*Infectious disease= HIV, AIDS, Coxsackievirus, Streptococcus pneumoniae, Parvovirus, Hepatitis C virus, Adenovirus, Chagas' disease, Lyme disease
\*\* Inflammatory disease= Rheumatic fever, Rheumatic disease, Sarcoidosis, Celiac disease
AM= acute myocarditis; VA=ventricular arrhythmia; ICD = implantable cardioverter defibrillator; NE- national estimate (see main text); CVA = cerebrovascular accident; TIA = transient ischemic attack

**Table 2:** Baseline characteristics acute myocarditis patients complicated by VT stratified by ablation status.

| Variable | Total Patients with<br>myocarditis and VT<br>(NE=1385) | Ablation<br>(NE=190) | No Ablation<br>(NE=1195) | p value |
| --- | --- | --- | --- | --- |
| Age (years) | 35 (19-57) | 31 (15-58) | 35 (19-57) | 0.207 |
| Female | 445 (32.1%) | 60 (31.6%) | 385 (32.2%) | 0.861 |
| White ethnicity | 780 (59.3%) | 80 (45.7%) | 700 (61.4%) | <0.001 |
| Diabetes mellitus | 65 (4.7%) | ≤10 | ≤10 | N/A |
| Hypertension | 425 (30.7%) | 55 (28.9%) | 370 (31.0%) | 0.576 |
| Dyslipidemia | 230 (16.6%) | 30 (15.8%) | 200 (16.7%) | 0.745 |
| Renal disease | 100 (7.2%) | ≤10 | ≤10 | N/A |
| Liver disease | 45 (3.2%) | ≤10 | ≤10 | N/A |
| Smoking | 350 (25.3%) | 30 (15.8%) | 320 (36.8%) | 0.001 |
| Obesity (BMI>30) | 100 (7.2%) | ≤10 | ≤10 | N/A |
| Valvular heart disease | 170 (12.3%) | 30 (15.8%) | 140 (5.3%) | 0.112 |
| Ischemic heart disease | 280 (20.2%) | 45 (23.7%) | 235 (19.7%) | 0.200 |
| Ischemic cardiomyopathy | 30 (2.2%) | ≤10 | ≤10 | N/A |
| Anemia | 55 (4%) | ≤10 | ≤10 | N/A |
| Alcohol abuse | 20 (1.4%) | ≤10 | ≤10 | N/A |
| Infectious disease* | 175 (12.6%) | 50 (26.3%) | 125 (10.5%) | <0.001 |
| Inflammatory disease** | 35 (2.5%) | ≤10 | ≤10 | N/A |
| CDMF-CCI score*** | 1 (0-2) | 1 (1-4) | 1 (0-2) | <0.001 |
| Income percentile |  |  |  |  |
| 0–25 | 325 (23.6%) | 20 (10.8%) | 305 (25.6%) | <0.001 |
| 26–50 | 265 (19.3%) | 30 (16.2%) | 235 (19.7%) |  |
| 51–75 | 390 (28.4%) | 90 (48.6%) | 300 (25.2%) |  |
| 76–100 | 395 (28.7%) | 45 (24.3%) | 350 (29.4%) |  |
| Hospital type |  |  |  |  |
| Rural | 35 (2.5%) | ≤10 | ≤10 | N/A |
| Urban, non-teaching | 160 (11.6%) | ≤10 | ≤10 |  |
| Urban, teaching | 1190 (85.9%) | ≤10 | ≤10 |  |

\*Infectious disease= HIV, AIDS, Coxsackievirus, Streptococcus pneumoniae, Parvovirus, Hepatitis C virus, Adenovirus, Chagas' disease, Lyme disease
\*\* Inflammatory disease= Rheumatic fever, Rheumatic disease, Sarcoidosis, Celiac disease
AM= acute myocarditis; NE- national estimate (see main text); VT=ventricular tachycardia

The Claims-based, Disease Specific refinements, Matching translation to ICD-9 and ICD-10, Flexibility Charlson Comorbidity Index (CDMF-CCI),^14,15^ a widely used adaptation of the Charlson Comorbidity Index, was calculated by identifying 17 weighted comorbidities based on ICD-10-CM codes available in the dataset. A detailed breakdown of the CDMF-CCI components is provided in Appendix Table 2. Higher scores on the CDMF-CCI reflect a greater burden of comorbid disease and are associated with increased risk of mortality within one year of hospitalization.^15–17^ This index has been extensively validated and is frequently applied in studies utilizing administrative datasets to predict both short- and long-term clinical outcomes.

The primary outcome of this study was a composite measure of in-hospital complications, encompassing arterial thromboembolic events (including cerebrovascular accident (CVA), transient ischemic attack (TIA), and systemic thromboembolism involving the lower or upper extremities, or the visceral-mesenteric circulation); cardiac complications [cardiogenic shock, cardiac arrest, vasopressor requirement, mechanical circulatory support, and pericardial complications (including pericardial tamponade, hemopericardium, pericarditis, and the need for pericardiocentesis)]. Additional components of the composite outcome included hemorrhagic events or hematomas, vascular injuries, requirement for mechanical ventilation, and all-cause in-hospital mortality.

Secondary outcomes consisted of the individual components of the primary outcome, implantable cardioverter defibrillator (ICD) implantations along with hospital length of stay and total hospitalization charges. Detailed definitions and corresponding ICD-10-CM/PCS codes for all outcomes are listed in Appendix Table 1.

### Statistical Analyses

Frequencies and proportions of demographic, clinical, and hospital-related variables were calculated and weighted to reflect national estimates using discharge sample weights provided by the NIS.

Comparative analyses were performed in two stages: First, among patients with acute myocarditis, comparing those who developed VA to those who did not; Multivariate analysis assessed the role of VA and other risk factors on in-hospital mortality, cardiac complications and total complications. Second, among the population of acute myocarditis complicated by VT, patients who underwent catheter ablation were compared to those who did not.

Categorical variables were compared using the chi-square (χ²) test. For continuous variables, we used the independent samples t-test when the distribution was normal, and the Mann–Whitney U test when normality assumptions were not met. Where appropriate, Bonferroni correction was applied to adjust for multiple comparisons.

All statistical analyses were performed using IBM SPSS Statistics, version 29. A p-value of <0.05 was considered statistically significant.

## RESULTS

### Study cohort

A total of 3,569 cases with a primary diagnosis of acute myocarditis were located within the database. After implementing the validated HCUP weighting method, these represented an estimated national total of 17,845 patients with acute myocarditis (Figure 1). The majority of patients were males (65.5%) and the mean age of the cohort was 35 (22-53) years. Within this cohort, we compared 1505 patients (8.4%) who developed VA to those who did not. A secondary analysis was performed among 1385 patients who developed VT (7.7%), comparing 190 patients who underwent catheter ablation (13.7% of VT patients) to those who did not.

### Patients’ characteristics

Baseline characteristics of the study population are presented in detail in Table 1. A total of 1,505 patients (8.4%) were diagnosed with VA. Patients who developed VA had significantly higher rates of renal and hepatic disease, atrial fibrillation, valvular heart disease, anemia, and infectious diseases, but not of inflammatory diseases. In addition, their overall comorbidity burden, as reflected by a higher CDMF-Charlson Comorbidity Index, was significantly higher. While the prevalence of ischemic heart disease was similar between the two groups, patients with VA had significantly higher rates of ischemic cardiomyopathy (p<0.001). Although age was not significantly different, the prevalence of diabetes, dyslipidemia, obesity, and smoking was significantly lower among myocarditis patients developing VA compared to those who did not.

Comparison of baseline characteristics between the 190 patients with acute myocarditis who underwent catheter ablation and those who did not (n=1385) is presented in Table 2. Among patients with VT, those who underwent ablation had significantly higher rates of atrial fibrillation (p=0.043) and infectious disease etiology (p<0.001). Although no statistically significant differences were observed in other underlying conditions, the CDMF-CCI score remained significantly higher in the ablation group (p<0.001).

### In-hospital course and outcomes

Patients with acute myocarditis who developed VA had a significantly higher rate of any in-hospital complications (50.5% vs. 15.7%, p<0.001). These included cardiac complications such as cardiogenic shock (30.9% vs. 5.8%, p<0.001), cardiac arrest (10% vs. 1.0%, p<0.001), vasopressor use (6.3% vs. 1.2%, p<0.001), and mechanical circulatory support (14.6% vs. 2.1%, p<0.001). Vascular complications were also more common in the VT group, including hemorrhage or hematoma (12.6% vs. 1.4%, p<0.001), as well as vascular injury (3% vs. 0.5%, p<0.001). The need for mechanical ventilation was higher (24.2% vs. 5.7%, p<0.001) and neurologic events, including stroke or transient ischemic attack (TIA), occurred more frequently (3.7% vs. 1.3%, p<0.001), as did the need for hemodialysis (2.3% vs. 0.7%, p<0.001). Furthermore, the VA group had significantly greater length of hospital stay, total hospital charges, and in-hospital mortality (10% vs. 1.6%, p<0.001)(Figure 2). In total, 115 patients (82% with VA) underwent ICD implantation during the index hospitalization. A complete summary of in-hospital outcomes of acute myocarditis by VA status is provided in Table 3.

**Figure 2:**
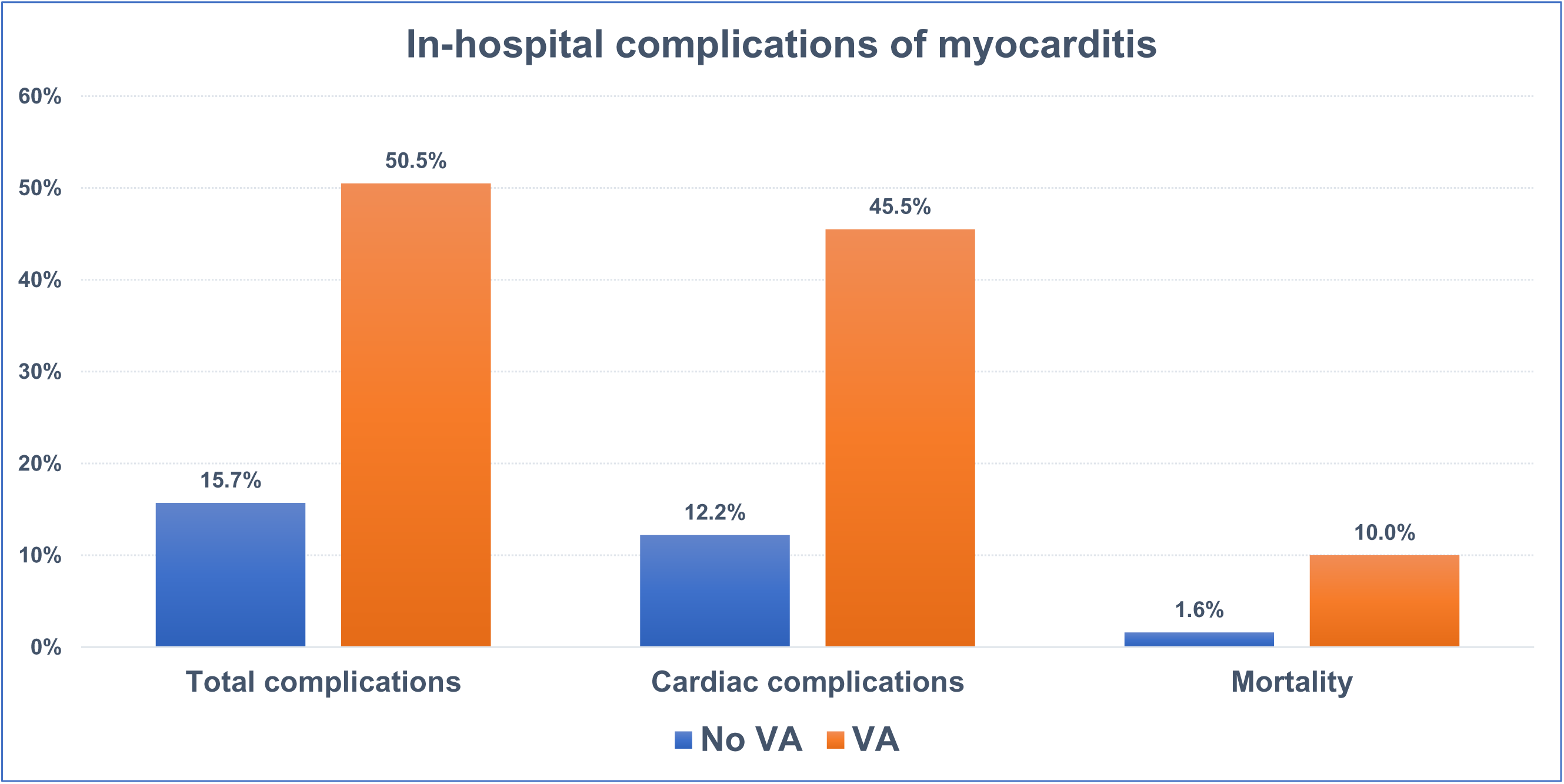
In hospital complications of myocarditis stratified by occurrence of VT.

**Table 3:** In-hospital outcomes of acute myocarditis patients complicated by VA.

| Variable | VA<br>(NE=1505) | No VA<br>(NE=16340) | p value |
| --- | --- | --- | --- |
| **Total complications** | 760 (50.5%) | 2570 (15.7%) | <0.001 |
| **Cardiac complications** | 685 (45.5%) | 1990 (12.2%) | <0.001 |
| Cardiogenic shock | 465 (30.9%) | 950 (5.8%) | <0.001 |
| Cardiac arrest | 150 (10%) | 160 (1.0%) | <0.001 |
| Pericardial complications* | 200 (13.3%) | 1135 (6.9%) | <0.001 |
| Vasopressor use | 95 (6.3%) | 200 (1.2%) | <0.001 |
| Mechanical circulatory support | 220 (14.6%) | 340 (2.1%) | <0.001 |
| **Noncardiac complications** |  |  |  |
| Hemorrhage/hematoma | 190 (12.6%) | 230 (1.4%) | <0.001 |
| Vascular injury | 45 (3.0%) | 80 (0.5%) | <0.001 |
| Mechanical ventilation | 335 (24.2%) | 945 (5.7%) | <0.001 |
| CVA/ TIA | 55 (3.7%) | 210 (1.3%) | <0.001 |
| Systemic embolism | 30 (2.0%) | 80 (0.5%) | <0.001 |
| Need for hemodialysis | 35 (2.3%) | 115 (0.7%) | <0.001 |
| **ICD implantation** | 95 (6.3%) | 20 (0.1%) | <0.001 |
| **Atrial fibrillation/flutter** | 210 (14.0%) | 940 (5.8%) | <0.001 |
| **Total hospital charges** | 92757 (43128-314018) | 35604 (21691-59593) | <0.001 |
| **Length of stay** | 5 (3-13) | 2 (1-4) | <0.001 |
| **In-hospital mortality** | 150 (10%) | 255 (1.6%) | <0.001 |

\* including tamponade, hemopericardium, pericarditis, pericardiocentesis
AM= acute myocarditis; VA=ventricular arrhythmia; ICD = implantable cardioverter defibrillator; NE- national estimate (see main text); CVA = cerebrovascular accident; TIA = transient ischemic attack

Multivariate analysis revealed that VA during admission for acute myocarditis was an independent significant risk factor for cardiac complications (aOR=4.8), total complications (aOR=4.2) and in hospital mortality (aOR=5.1) (p<0.001 for each analysis).

A subgroup analysis of the VT cohort (n=1385) compared 190 patients who underwent catheter ablation to those who did not (Table 4). The ablation group experienced markedly higher rates of cardiogenic shock (52.6% vs. 25.5%, p<0.001), cardiac arrest (13.2% vs. 4.2%, p<0.001), and required vasopressor support and mechanical circulatory assistance more frequently (10.5% vs. 5.4%, p=0.007; 39.5% vs. 10.9%, p<0.001, respectively). Vascular and respiratory complications were also significantly more prevalent among patients who underwent ablation, including hemorrhage or hematoma (31.6% vs. 9.6%, p<0.001) and mechanical ventilation (52.6% vs. 19.7%, p<0.001). Neurological events, such as stroke or TIA, occurred more frequently in this group (7.9% vs. 3.3%, p=0.003). In addition, patients in the ablation group had significantly longer hospitalizations, higher overall costs, and a greater rate of in-hospital mortality (15.8% vs. 6.7%, p<0.001).

**Table 4:**
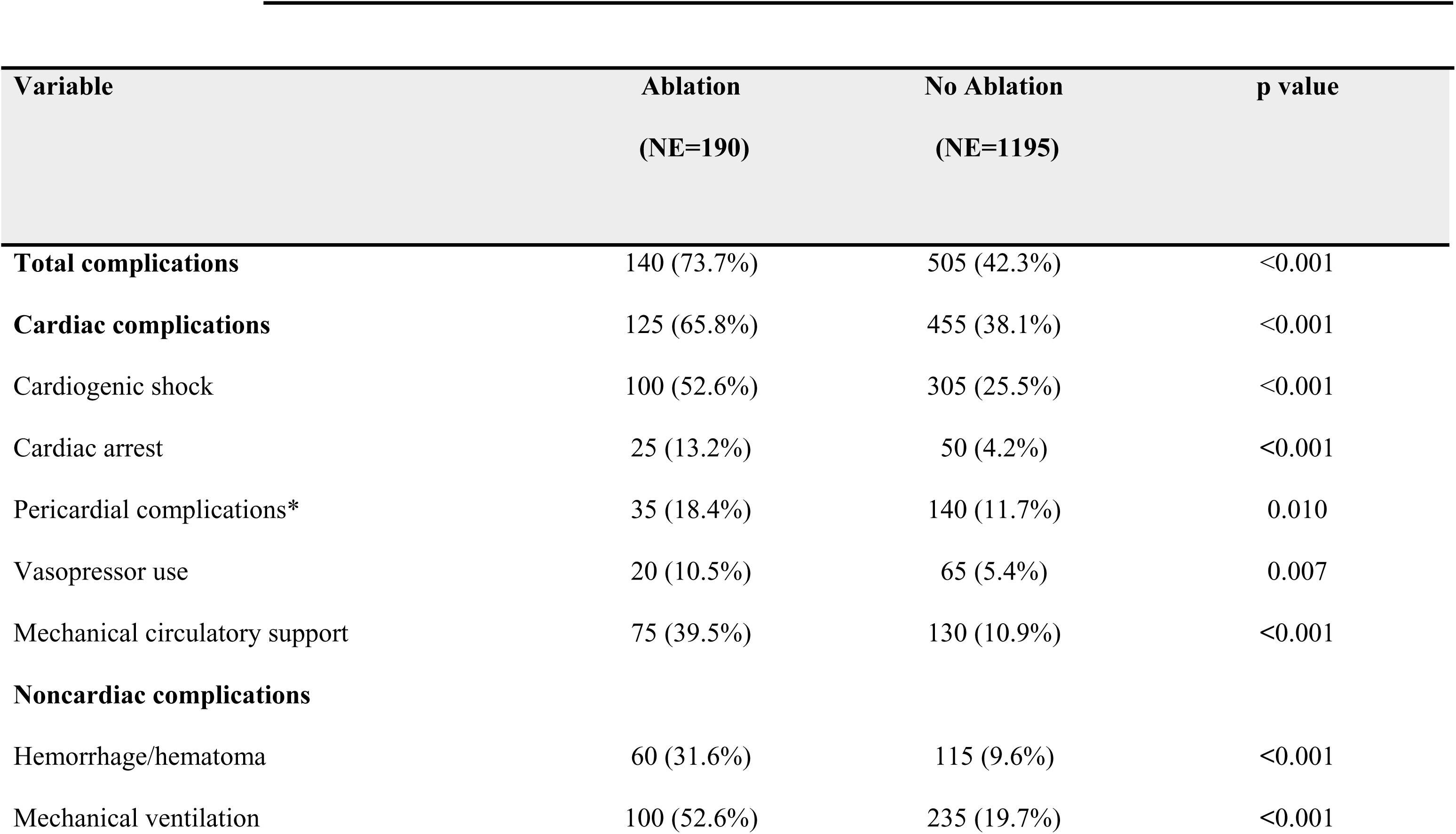
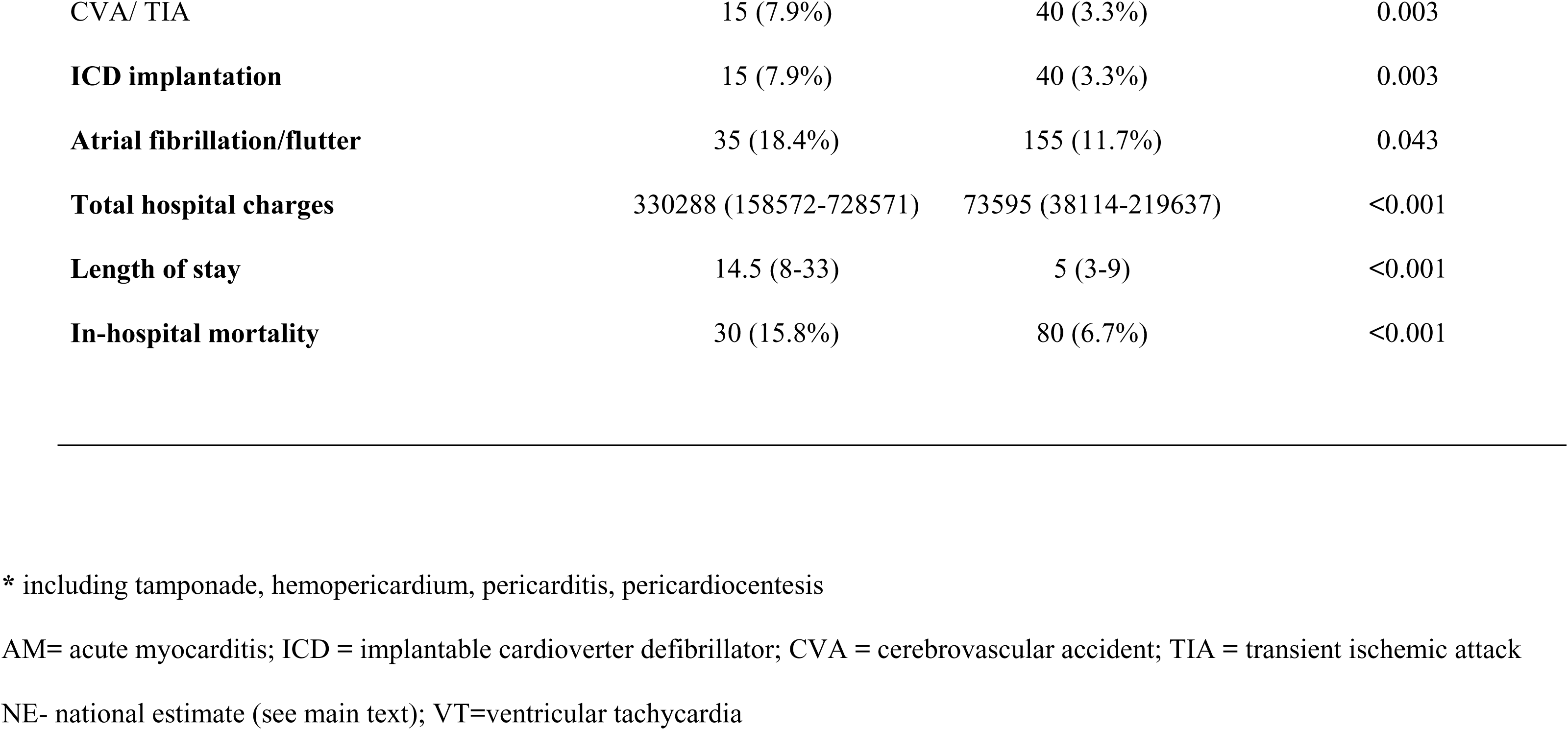
In-hospital outcomes of acute myocarditis patients complicated by VT stratified by ablation status.

## DISCUSSION

In this US nationwide analysis of 17,845 weighted hospitalizations for acute myocarditis between 2016 and 2019, we found that VA occurred in 8.4% of patients (with 7.7% having VT). Patients who developed VA had substantially greater in-hospital morbidity and mortality, including more cardiogenic shock, cardiac arrest, need for mechanical circulatory support, and a more than five-fold higher in-hospital mortality compared with patients without VA. Within the VT subgroup, 13.7% underwent catheter ablation during the index admission. These patients, selected for catheter ablation at the discretion of treating physicians, demonstrated markedly worse outcomes than their non-ablated counterparts, reflecting either the severity of the underlying inflammatory substrate, the hemodynamic compromise prompting intervention, or the procedural risks inherent to catheter ablation in an acutely inflamed myocardium.

Prior studies have shown that VA complicating acute myocarditis is associated with greater comorbidity burden and poorer in-hospital outcomes.^1,3^ Our results align with these findings, identifying acute myocarditis associated VA as a marker of a high-risk subgroup characterized by significantly higher comorbidity burden, more frequent systemic involvement (including renal dysfunction and anemia), and markedly worse in-hospital outcomes. Interestingly, traditional atherosclerotic disease risk factors including diabetes, dyslipidemia, obesity, and smoking, were less frequent among VT patients, supporting the concept that myocarditis driven arrhythmogenesis is mediated more by the inflammatory substrate.^1^

In our study, catheter ablation during the index hospitalization was performed in 1 out of 7 acute myocarditis patients with VT. Those who underwent ablation exhibited a higher prevalence of infection, atrial arrhythmias, and elevated comorbidity scores, along with markedly greater rates of cardiogenic shock (52%), cardiac arrest, requirement for mechanical circulatory support, mechanical ventilation, and bleeding complications. Due to limitations inherent in the NIS database, we cannot definitively determine whether these comorbidities and complications prompted referral for ablation or resulted from the procedure itself. Nevertheless, these findings indicate that ablation was predominantly performed in the most critically ill patients with VT in the setting of acute myocarditis.

Importantly, in-hospital mortality was elevated in the ablation group (15.8%), which may reflect the greater severity of their underlying condition rather than ablation failure per se. This interpretation aligns with prior data showing that bailout ablation for refractory ventricular tachycardia (VT) in the setting of cardiogenic shock is associated with an in-hospital mortality rate of 29%.^18^ Nevertheless, VT ablation during acute myocarditis was recently associated with increased risk of VT recurrence in two small-scale studies. ^9,10^ Taken together, our study findings highlight the need for further studies to clarify the role, timing, and outcomes of ablation in acute myocarditis associated VA.

## LIMITATIONS

Several important limitations should be addressed. Our study, based on the NIS database is a retrospective administrative dataset relying on ICD-10 coding, which may introduce misclassification of myocarditis etiology, arrhythmia type, and procedural details. Myocarditis subtypes (viral, autoimmune, giant-cell, eosinophilic) differ substantially in arrhythmic risk, but cannot be fully differentiated from ICD codes alone. In addition, the database lacks clinical granularity for key clinical variables including CMR findings, ejection fraction, histological results, biomarker levels, ECG patterns, VT burden, VT mechanism and duration, hemodynamic parameters, and medication use. Selection bias should be acknowledged regarding the decision whether to perform catheter ablation in that sub-population, and those results need to be considered in that context. Finally, the NIS database is limited to in-hospital data and lacks long-term follow-up, precluding assessment of post-discharge outcomes such as long-term mortality, ventricular arrhythmia recurrence, or readmission rates.

## CONCLUSIONS

In this large national cohort, VA occurred in nearly 8.5% of patients hospitalized with acute myocarditis and was strongly associated with increased morbidity and more than five-fold higher in-hospital mortality. Catheter ablation was performed in approximately 14% of myocarditis patients with VT and was associated with significantly worse in-hospital outcomes, compared to those who did not have ablation, likely reflecting both the severity of underlying disease and the risks of performing ablation in an acutely inflamed myocardium. These findings underscore the need for improved risk stratification, careful patient selection, and prospective studies to determine the optimal timing and role of catheter ablation in the management of acute myocarditis associated VA.

## Data Availability

All raw data can be obtained from the HCUP, part of the AHRQ, a federal agency.

## Acknowledgement

This manuscript is submitted in partial fulfillment of the requirements for the MD degree of Tzurit Kusniec at the Rappaport Faculty of Medicine, Technion – Israel Institute of Technology.

